# Using deep learning to predict brain age from brain magnetic resonance images and cognitive tests reveals that anatomical and functional brain aging are phenotypically and genetically distinct

**DOI:** 10.1101/2021.06.22.21259280

**Authors:** Alan Le Goallec, Samuel Diai, Sasha Collin, Théo Vincent, Chirag J. Patel

## Abstract

With the world population aging, the prevalence of age-related brain diseases such as Alzheimer’s, Parkinson’s, Lou Gehrig’s, and cerebrovascular diseases. In the following, we built brain age predictors by leveraging 46,000 brain magnetic resonance images [MRIs] and cognitive tests from UK Biobank participants. We predicted age with a R-Squared [R^2^] of 76.4±1.0% and a root mean squared error of 3.58±0.05 years and identified the features driving the prediction using attention maps. We defined accelerated brain aging as the difference between brain age (predicted age) and age. Accelerated brain aging is partially heritable (h_g^2^=35.9±2.6%), and is associated with 219 single nucleotide polymorphisms [SNPs] in 25 genes (e.g CRHR1, involved in the hypothalamic-pituitary-adrenal pathway). Similarly, it is associated with biomarkers (e.g blood pressure), clinical phenotypes (e.g general health), diseases (e.g diabetes), environmental (e.g smoking) and socioeconomic variables (e.g income and education). We performed the same analysis, this time distinguishing between anatomical (MRI-based) and functional (cognitive tests-based) brain aging. We found the two accelerated aging phenotypes to be phenotypically .112±.006 correlated and genetically uncorrelated, with distinct SNPs and non-genetic factors associated with each. In conclusion, anatomical and functional brain aging are two distinct, complex phenotypes, which also differ in their genetic and non-genetic factors. Our brain predictors could be used to monitor the effects of emerging rejuvenating therapies on the brain.

## Background

As we age, the brain undergoes important changes, both anatomically (atrophy, white matter hyperintensities, impaired white matter integrity, microbleeds, small vessel changes) ^1^ and functionally (reduced processing speed and fluid intelligence) ^2, 3^. The brain also becomes more likely to develop age-related diseases such as Alzheimer’s, Parkinson’s, Lou GeHrig’s and cerebrovascular diseases ^4^. With the world population aging ^5^, it is expected that the prevalence of these deadly and incapacitating diseases will increase.

To better understand brain aging, brain age predictors can be built by training machine learning models to predict age. The prediction for an unseen sample can be interpreted as the participant’s brain age. Accelerated brain aging is defined as the difference between brain age and age (also referred to as chronological age, to distinguish it from biological age, here brain age). A large number of brain age predictors have been built from brain MRI images ^6–22^ and electroencephalograms ^23, 24^. To our knowledge, only one age predictor was built on cognitive tests, on a limited sample size (N=537) ^25^. Additionally, while brain anatomy and function are linked (e.g changes in processing speed are associated with changes in the cerebellar and the prefrontal cortex ^26^), the connection between anatomical (MRI-based) and functional (cognitive tests-based) accelerated brain aging has not been investigated.

In the following, we leverage 46,000 brain magnetic resonance images [MRIs] (Figure 1B) and cognitive tests from UK Biobank ^27^ participants aged 37-82 year-old to build brain age predictors capturing different facets of brain anatomical and functional aging. We then hierarchically ensembled these predictors into three brain age predictors: brain general age, brain anatomical age and brain cognitive age (Figure 1A). We performed a genome wide association study to identify single nucleotide polymorphisms [SNPs] associated with accelerated aging in each brain dimension. Similarly, we performed an X-wide association study to identify biomarkers, clinical phenotypes, diseases, environmental and socioeconomic variables associated with each accelerated brain aging dimension. Finally, we computed the phenotypic, genetic and non-genetic correlation between accelerated brain anatomical and cognitive aging. (Figure 1C)

**Figure 1:**
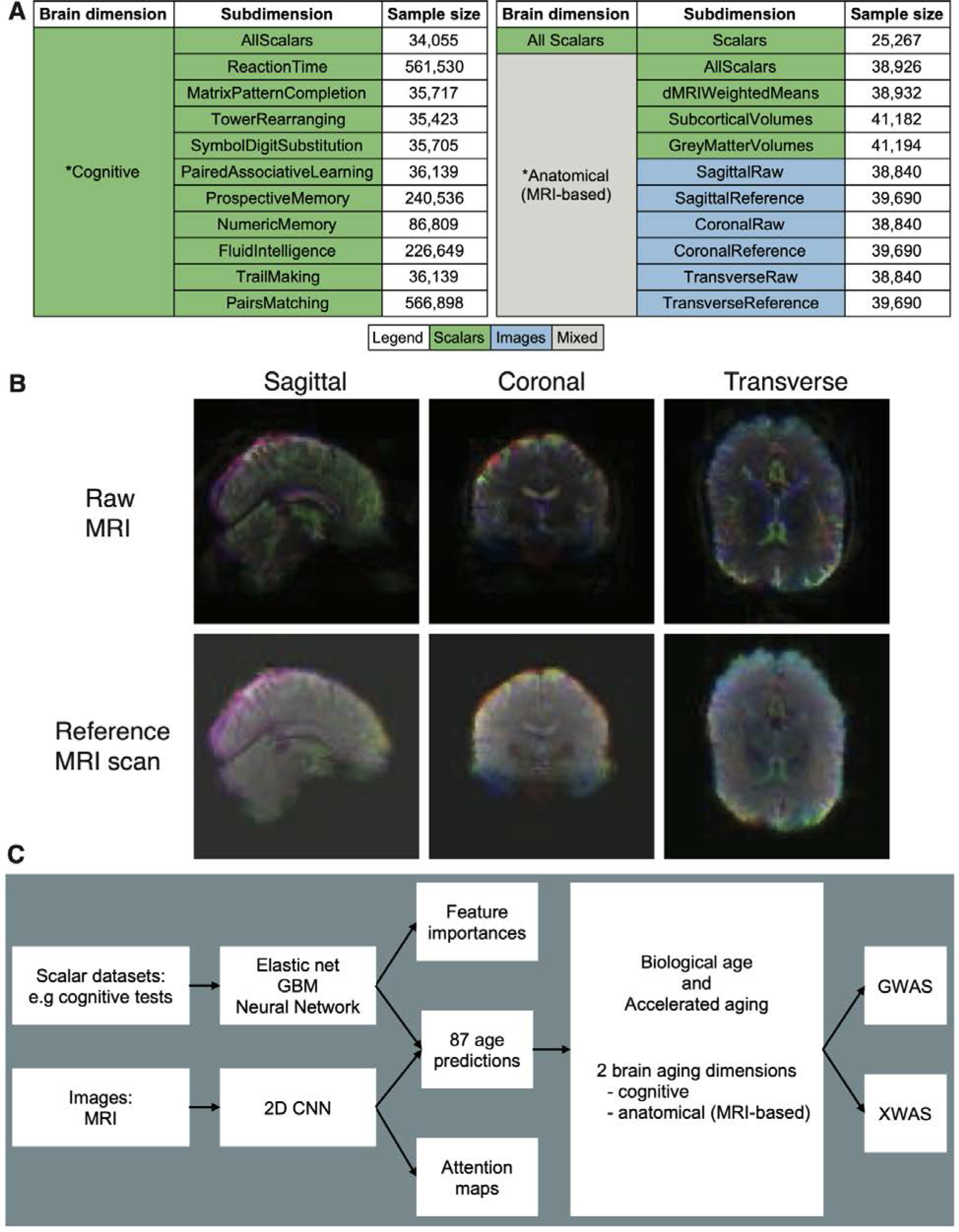
Overview of the datasets and analytic pipeline. A - Brain aging dimensions and subdimensions. B - Brain MRI samples. C - Analytic pipeline. Legend for C - GWAS: Genome Wide Association Study. XWAS: X-Wide Association Study.

## Results

### We predicted age within four years

We leveraged the UK Biobank, a dataset collected of 502,211 participants with 46,000 three-dimensional brain MRI videos and cognitive tests (Figure 1A). For each three-dimensional video, we extracted a sagittal, coronal and transverse view image of the median time frame, and did the same for the “Reference scan” (Figure 1B). We predicted age from images using convolutional neural networks [CNN] and from scalar features using elastic nets, gradient boosted machines and shallow, fully connected neural networks. We then hierarchically ensembled the models (Figure 1A) to obtain a prediction for each brain age dimension and subdimension (Figure 2). The all-encompassing ensemble model predicted age with a R^2^ of 76.4±1.0% and a RMSE of 3.58±0.05 years. Age was predicted significantly better by anatomical features than by cognitive tests (R^2^=75.3±0.8% vs. 37.9±1.1%). For the models built on MRI data, the ensemble model encompassing all data modalities (images and scalars) significantly outperformed the ensemble model that relied exclusively on scalar features (R^2^=75.3±0.8% vs. 64.7±1.0%). For scalar-based models, non-linear algorithms (GBM, neural network) only marginally outperformed linear models (cognitive tests: R^2^=37.8±1.2% vs. 34.0±1.4%; MRI features: R^2^=62.7±1.7% vs. 62.2±1.1%).

**Figure 2:**
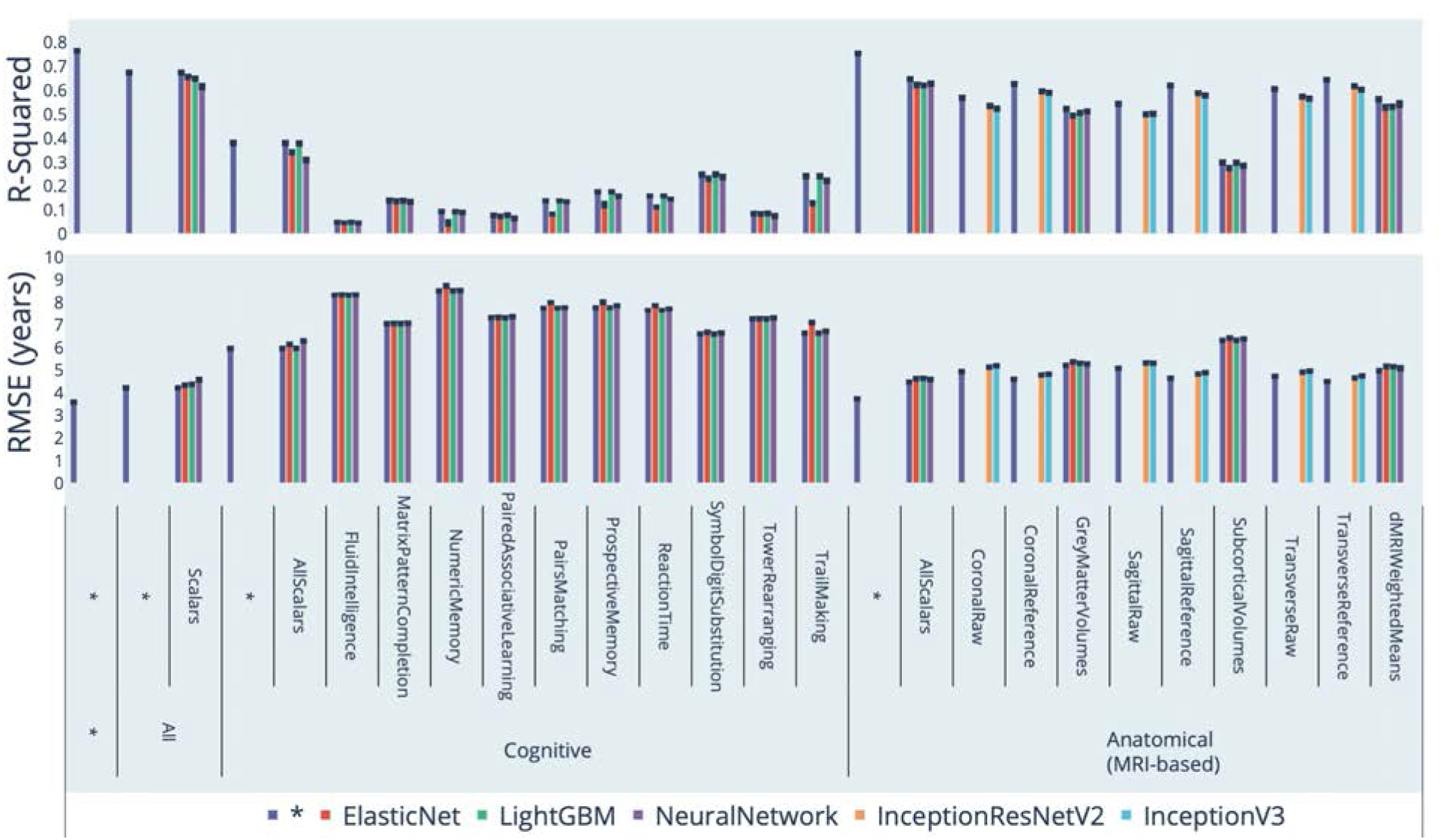
Chronological age prediction performance (R^2^ and RMSE) * represent ensemble models

### Correlation between anatomical and cognitive brain aging

We defined brain age as the prediction outputted by a model trained to predict brain age, after correction for the bias in the residuals (see methods). The predictors trained on different dimensions and subdimensions of brain aging (Figure 1A) yielded different definitions of brain aging (e.g anatomical brain aging vs. cognitive brain aging). We defined accelerated aging phenotypes as the difference between brain age and chronological age (commonly referred to as “age”, the time since an individual’s birth). We then computed the correlation between accelerated aging in the different brain dimensions and subdimensions (Figure 3). We found that anatomical and cognitive accelerated brain aging are weakly correlated (.112±.006). Accelerated aging in subdimensions under the same brain dimension tend to be more correlated. For example, the brain cognitive sub-subdimensions (reaction time, matrix pattern completion, tower rearranging, symbol digit substitution, paired associative learning, prospective memory, numeric memory, fluid intelligence, trail making and paris matching) are .391±0.098 correlated (based on 45 pairwise correlations), and brain MRI image-based anatomical subdimensions are .448±.072 correlated (based on 18 pairwise correlations).

**Figure 3:**
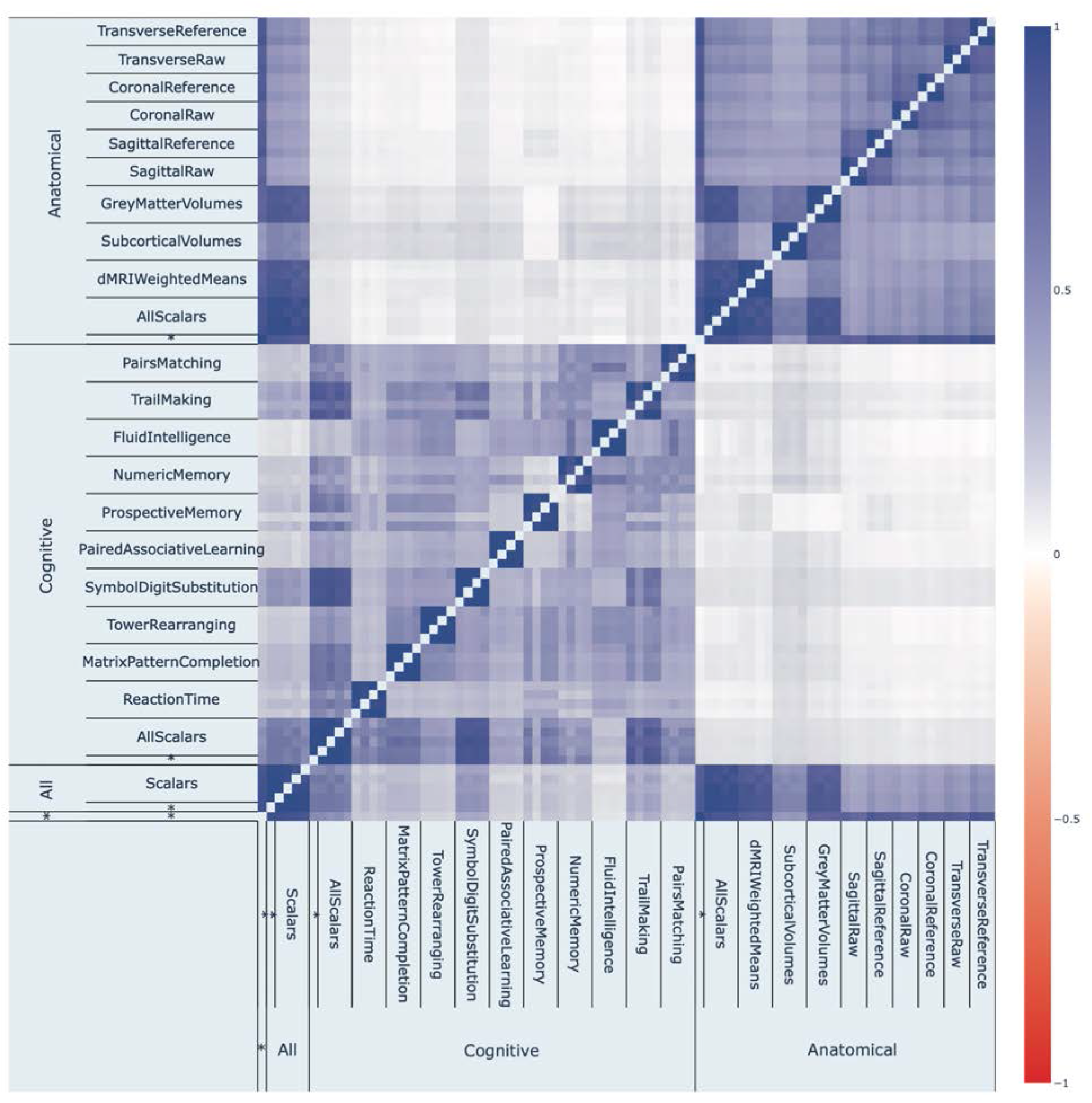
**Phenotypic correlation between accelerated aging for the different brain dimensions and subdimensions**

**Figure 4:**
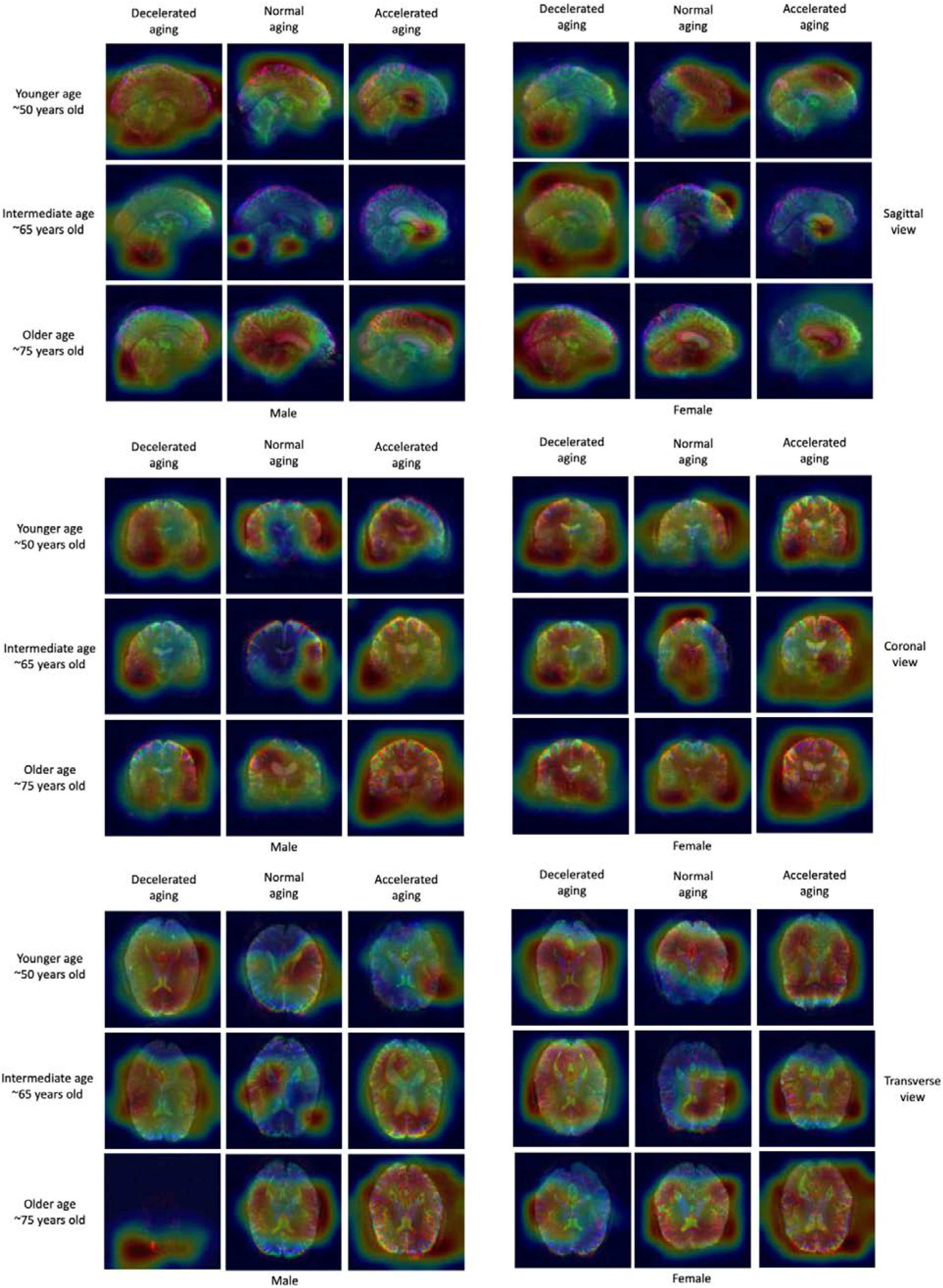
Attention map samples for MRI-based models. Warm colors highlight regions of high importance according to the Grad-RAM map. All images are reference scans collected from 60-65 year-old females.

### Identification of the anatomical and cognitive features driving brain age prediction

We best predicted brain cognitive age with a gradient boosted machine that gave a R^2^ of 37.8±1.2% (R^2^=37.9±0.3% for the ensemble model based on all cognitive tests). The model was trained on 135 biological features, along with sex and ethnicity. We report the feature importances in Table S2. We found that the nine most predictive scalar features are all related to speed of answering questions and include, in order: (1) time to answer, (2) mean time to correctly identify matches, (3) duration to complete alphanumeric path (trail #2), (4) time to complete test, (5) duration to complete numeric path (trail #1), (6) time to complete round (second round), (7) time to complete round (first round), (8) mean time to solve puzzles and (9) mean time to press snap-button. These nine features were associated with older age based on their elastic net (R^2^=33.0) regression coefficients. The tenth most important feature selected by the GBM was the fluid intelligence score, which is negatively associated with aging based on the elastic net regression coefficient. The fluid intelligence score (UKB field 20,016) is the number of correct answers to 13 questions: numerical addition test, identify largest number, word interpolation, positional arithmetic, family relationship calculation, conditional arithmetic, synonym, chained arithmetic, concept interpolation, arithmetic sequence recognition, antonym, square sequence recognition and subset inclusion logic.

In terms of scalar features, we best predicted brain anatomical aging using a neural network (R^2^=62.7±1.7%). The elastic net (R^2^=62.2±1.1%) and the GBM (R^2^=62.0±1.0%) performed similarly well. The models were trained on 396 biological features, along with sex and ethnicity. We report the feature importances in Table S3. The neural network highlighted (1) the forceps minor, (2) the forceps major, (3; 4) the right superior longitudinal fasciculus, (5; 7) the left acoustic radiation, (6) the right inferior longitudinal fasciculus, (8) the left corticospinal tract and (9; 10) the right and left inferior fronto-occipital fasciculi. The GBM highlighted (1) the volume of gray matter in the thalamus (right side), (2; 10) the posterior thalamic radiations (right and left sides), (3) the volume of gray matter in the ventral striatum (left side), (4) the forceps minor, (5) the superior thalamic radiation (right side), (6) the corticospinal tract (left side), (7) the subcallosal cortex (right side), (8) the volume of the putamen (left side) and (9) the volume of gray matter in the insular cortex (right side). Based on the elastic net regression coefficients, the five features most associated with old age are (1) the anterior thalamic radiation, (2) the acoustic radiation, (3) the inferior longitudinal fasciculus, (4) the volume of gray matter in the subcallosal cortex and (5) the posterior thalamic radiation (all on the right side). The five features most associated with young age are (1) the acoustic radiation (left side), (2) the corticospinal tract (left side), (3) the forceps minor, (4) the ventral striatum (left side) and (5) the middle cerebellar peduncle.

We report the correlation between the feature importances estimated by the different algorithms in Table S4 and Table S5.

In terms of brain MRI images, the attention maps highlighted different features across participants (Figure S15).

### Genetic factors and heritability of accelerated brain aging

We performed three genome wide association studies [GWASs] to estimate the GWAS-based heritability of general (h_g^2^=34.9±1.8%), anatomical (h_g^2^=30.7±1.6%), and cognitive (h_g^2^=32.9±1.7%) accelerated brain aging, and to identify SNPs associated with these phenotypes (Figure 5 and Table 1). Accelerated anatomical and cognitive brain aging are genetically uncorrelated (correlation=.019±.056).

**Figure 5:**
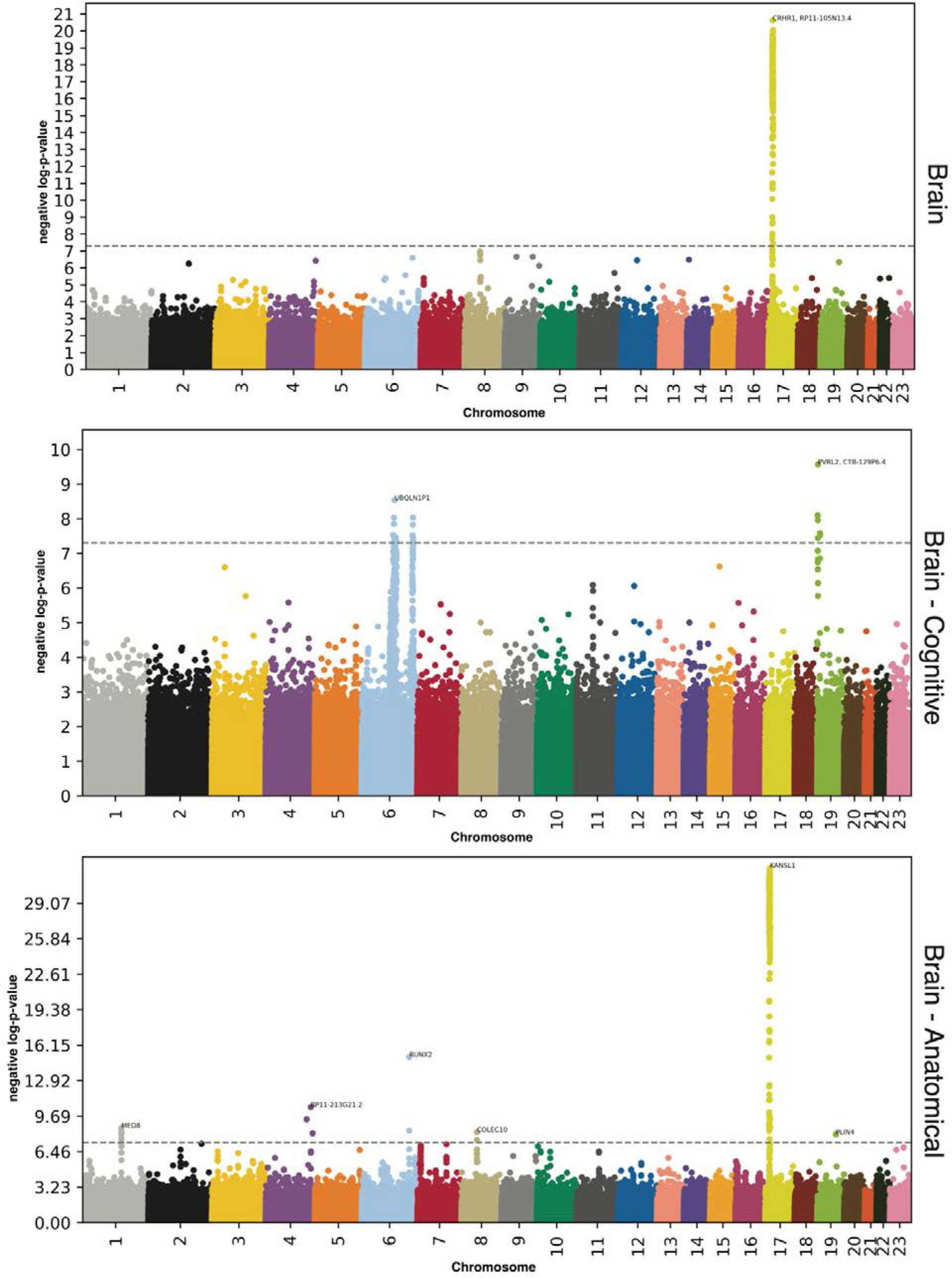
GWAS results - SNPs associated with accelerated brain aging for each brain dimension. -log10(p-value) vs. chromosomal position of locus. Dotted line denotes 5x10^-8^.

**Table 1:** GWASs summary - Heritability, number of SNPs and genes associated with accelerated aging in each brain dimension.

| Brain dimension | Sample size | SNPs | Genes | Heritability (%) | R-Squared for CA prediction (%) |
| --- | --- | --- | --- | --- | --- |
| General | 22,951 | 219 | 25 | 35.9±2.6 | 76.4±1.0 |
| Cognitive | 30,108 | 27 | 20 | 29.3±2.0 | 37.9±1.1 |
| Anatomical | 35,351 | 235 | 35 | 39.1±1.7 | 75.3±0.8 |

#### General brain aging

We found that accelerated brain aging is 35.9±2.6% heritable and that 219 SNPs in 25 genes are significantly associated with this phenotype. The GWAS highlighted a single peak with the gene CRHR1 (Corticotropin Releasing Hormone Receptor 1, a GPCR protein involved in the hypothalamic-pituitary-adrenal pathway). CRHR1 is in linkage disequilibrium with other genes such as CRHR1-IT1, (CRHR1 intronic transcript 1), RP11-105N13.4 and RP11-669E14.4 (a processed transcript with unknown functions and an antisense gene linked to Alzheimer’s disease, both in the Pre-MRNA Processing Factor 31 [PRPF31] gene. PRPF31 is a component of the spliceosome complex and is linked to Retinitis Pigmentosa) and LRRC37A2 (Leucine Rich Repeat Containing 37 Member A2, linked to progressive myoclonic epilepsy-6 and Koolen-De Vries syndrome).

#### Anatomical brain aging

We found that accelerated anatomical (MRI-based) brain aging is 39.1±1.7% heritable and that 235 SNPs in 35 genes are significantly associated with this phenotype. The GWAS highlighted six peaks: (1) KANSL1 (KAT8 Regulatory NSL Complex Subunit 1, involved in chromatin organization through histone acetylation and linked to Koolen-De Vries Syndrome). KANSL1 is in linkage disequilibrium with other genes such as MAPT (Microtubule Associated Protein Tau, linked to diverse neurodegenerative disorders such as frontotemporal dementia, progressive supranuclear palsy, Alzheimer’s disease, Pick’s disease and cortico-basal degeneration), MAPT-AS1 (MAPT Antisense RNA 1, a long non-coding RNA linked to diseases such as late onset Parkinson Disease and Koolen-De Vries Syndrome) and CRHR1 (Corticotropin Releasing Hormone Receptor 1, a GPCR protein involved in the hypothalamic-pituitary-adrenal pathway); (2) RUNX2 (RUNX Family Transcription Factor 2, involved in osteoblastic differentiation and skeletal morphogenesis); (3) RP11-213G21.2 (a long intergenic non-coding RNA). RP11-213G21.2 is in linkage disequilibrium with RPS23P3 (Ribosomal Protein S23 Pseudogene 3, involved in RNA synthesis); (4) MED8 (Mediator Complex Subunit 8, involved in RNA synthesis); (5) COLEC10 (Collectin Subfamily Member 10, a C-lectin cytosolic protein); and (6) PLIN4 (Perilipin 4, involved in lipid storage).

#### Cognitive brain aging

We found that accelerated cognitive aging is 29.3±2.0% heritable and that 27 SNPs in 20 genes are significantly associated with this phenotype. The GWAS highlighted three peaks: (1) PVRL2 (Poliovirus Receptor-Related 2 (Herpesvirus Entry Mediator B), involved in Herpes cell to cell transmission). PVRL2 is in linkage disequilibrium with CTB-129P6.4 (an antisense gene), TOMM40 (Translocase Of Outer Mitochondrial Membrane 40, involved in the transport of protein precursors into mitochondria) and APOC1 (Apolipoprotein C1, involved in high density lipoprotein (HDL) and very low density lipoprotein (VLDL) metabolism); (2) UBQLN1P1 (Ubiquilin 1 Pseudogene 1, a pseudogene). UBQLN1P1 is in linkage disequilibrium with several genes such as GNL1 (G Protein Nucleolar 1, part of the major histocompatibility complex), ATAT1 (Alpha Tubulin Acetyltransferase 1, involved in microtubule dynamics), C6orf136 (Chromosome 6 Open Reading Frame 136, linked to Central Nervous System Germ Cell Tumor and Spinocerebellar Ataxia), PPP1R18 (Protein Phosphatase 1 Regulatory Subunit 18, linked to deafness), MDC1 (Mediator Of DNA Damage Checkpoint 1, involved in DNA damage response); and (3) UBDP1 (Ubiquitin D Pseudogene 1, a pseudogene). UBDP1 is in linkage disequilibrium with OR5V1 (Olfactory Receptor Family 5 Subfamily V Member 1, involved in olfactory transduction) and XXbac-BPG13B8.10 (a long intergenic non-coding RNA).

### Biomarkers, clinical phenotypes, diseases, environmental and socioeconomic variables associated with accelerated brain aging

We use “X” to refer to all nongenetic variables measured in the UK Biobank (biomarkers, clinical phenotypes, diseases, family history, environmental and socioeconomic variables). We performed an X-Wide Association Study [XWAS] to identify which of the 4,372 biomarkers classified in 21 subcategories (Table S6), 187 clinical phenotypes classified in 11 subcategories (Table S9), 2,073 diseases classified in 26 subcategories (Table S12), 92 family history variables (Table S15), 265 environmental variables classified in nine categories (Table S18), and 91 socioeconomic variables classified in five categories (Table S21) are associated (p-value threshold of 0.05 and Bonferroni correction) with accelerated brain aging in the different brain dimensions. We summarize our findings for general accelerated anatomical aging and cognitive aging in Table S7, Table S8, Table S10, Table S11, Table S13, Table S14, Table S19, Table S20, Table S22, Table S23, where we list the three variables of each X-subcategory (e.g spirometry biomarkers) most associated with each accelerated brain aging dimension. The full results can be exhaustively explored at https://www.multidimensionality-of-aging.net/xwas/univariate_associations.

#### Biomarkers associated with accelerated brain aging

##### Anatomical aging

The three biomarker categories most associated with accelerated brain anatomical aging are urine biochemistry, blood pressure, and trail making (a cognitive test). Specifically, 100.0% of urine biochemistry biomarkers are associated with accelerated brain anatomical aging, with the three largest associations being with microalbumin (correlation=.050), sodium (correlation=.038) and creatinine (correlation=.029). 100.0% of blood pressure biomarkers are associated with accelerated brain anatomical aging, with the three largest associations being with systolic blood pressure (correlation=.079), diastolic blood pressure (correlation=.075), and pulse rate (correlation=.060). 75.0% of trail making biomarkers are associated with accelerated brain anatomical aging, with the three largest associations being with the duration to complete an alphanumeric path (correlation=.048), the duration to complete a numeric path (correlation=.044), and the total errors traversing an alphanumeric path (correlation=.033).

Conversely, the three biomarker categories most associated with decelerated brain anatomical aging are hand grip strength, spirometry and symbol digit substitution (a cognitive test). Specifically, 100.0% of hand grip strength biomarkers are associated with decelerated brain anatomical aging, with the two associations being with left and right hand grip strengths (correlations of respectively .076 and .072). 100.0% of spirometry biomarkers are associated with decelerated brain anatomical aging, with the three largest associations being with forced vital capacity (correlation=.106), forced expiratory volume in one second (correlation=.104), and peak expiratory flow (correlation=.066). 100.0% of symbol digit substitution biomarkers are associated with decelerated brain anatomical aging, with the two associations being with the number of symbol digits matches made correctly (correlation=.106) and the number of digit matches attempted (correlation=.104).

##### Cognitive aging

The three biomarker categories most associated with accelerated cognitive aging, aside from the cognitive tests on which the models were built, are hearing, brain MRI weighted means and body impedance. Specifically, 60.3% of hearing biomarkers are associated with accelerated cognitive aging, with the three largest associations being with the duration of the hearing test on the right and left sides (respective correlations of .127 and .122), and the time to press next (correlation=.109). 41.2% of brain MRI weighted means biomarkers are associated with accelerated cognitive aging, with the three largest associations being with features in the right tract posterior thalamic radiation (correlations of .093 and .091), and left anterior thalamic radiation (correlation=.088). 40.0% of body impedance biomarkers are associated with accelerated cognitive aging, with the two associations being with right and left arm impedances (respective correlations of .038 and .030).

Conversely, the three biomarker categories most associated with decelerated cognitive aging are hand grip strength, spirometry and brain subcortical volumes. Specifically, 100.0% of hand grip strength biomarkers are associated with decelerated cognitive aging, with the two associations being with left and right hand grip strength (respective correlations of .062 and .057). 100.0% of spirometry biomarkers are associated with decelerated cognitive aging, with the three largest associations being with peak expiratory flow (correlation=.081), forced expiratory volume in one second (correlation=.078), and forced vital capacity (correlation=.071). 85.7% of brain subcortical volumes are associated with decelerated brain aging, with the three largest associations being with the volumes of the thalamus on the left and right sides (respective correlations of .108 and .106) and the volume of the putamen on the left side (correlation=.075).

#### Clinical phenotypes associated with accelerated brain aging

##### Anatomical aging

The three clinical phenotype categories most associated with accelerated brain anatomical aging are breathing, chest pain and general health. Specifically, 100.0% of breathing phenotypes are associated with accelerated brain anatomical aging, with the two associations being with shortness of breath walking on level ground (correlation=.068) and wheezing or whistling in the chest in the last year (correlation=.034). 75.0% of chest pain phenotypes are associated with accelerated brain anatomical aging, with the three associations being with chest pain due to walking ceases when standing still (correlation=.033), chest pain or discomfort when walking uphill or hurrying (correlation=.025), and chest pain or discomfort walking normally (correlation=.024). 50.0% of general health phenotypes are associated with accelerated brain anatomical aging, with the three largest associations being with overall health rating (correlation=.095), long-standing illness, disability or infirmity (correlation=.079), and falls in the last year (correlation=.037).

Conversely, the three clinical phenotype categories most associated with decelerated brain anatomical aging are sexual factors (age first had sexual intercourse: correlation=.029), mouth health (no mouth/teeth problems: correlation=.037) and general health (no weight change during the last year: correlation=.034).

##### Cognitive aging

The three clinical phenotype categories most associated with accelerated cognitive aging are breathing, chest pain, and mental health. Specifically, 100.0% of breathing phenotypes are associated with accelerated brain anatomical aging, with the two associations being with shortness of breath walking on level ground (correlation=.056) and wheezing or whistling in the chest in the last year (correlation=.036). 50.0% of chest pain phenotypes are associated with accelerated cognitive aging, with the two associations being with chest pain or discomfort walking normally (correlation=.031) and chest pain or discomfort (correlation=.027). 41.3% of mental health phenotypes are associated with accelerated cognitive aging, with the three largest associations being with nervous feelings (correlation=.067), worrier/anxious feelings (correlation=.065), and frequency of tiredness/lethargy in the last two weeks (correlation=.056).

Conversely, the three clinical phenotype categories most associated with decelerated cognitive aging are mouth health (no bucal/dental problem: correlation=.028), general pain (no pain experienced in the last month: correlation=.032) and mental health (no manic/hyper syndrome and no manic/irritable episodes: respective correlations of .066 and .058).

#### Diseases associated with accelerated brain aging

##### Anatomical aging

The three disease categories most associated with accelerated brain anatomical aging are metabolic, cardiovascular and mental disorders. Specifically, 10.1% of metabolic diseases are associated with accelerated brain anatomical aging, with the three largest associations being with non-insulin-dependent, insulin-dependent and unspecified type two diabetes (respective correlations of .089, .054 and .041). 9.1% of cardiovascular diseases are associated with accelerated brain anatomical aging, with the three largest associations being with hypertension (correlation=.082), chronic ischaemic heart disease (correlation=.039), and acute myocardial infarction (correlation=.031). 5.4% of mental disorders are associated with accelerated brain anatomical aging, with the three largest associations being with mental and behavioral due to use of alcohol or tobacco (respective correlations of .049 and .039), and depressive episode (correlation=.032).

We found no disease to be associated with accelerated brain anatomical aging.

##### Cognitive aging

The three disease categories most associated with accelerated cognitive aging are cardiovascular, general health and metabolic diseases. Specifically, 3.9% of cardiovascular diseases are associated with accelerated cognitive aging, with the three largest associations being with hypertension (correlation=.036), chronic ischaemic heart disease (correlation=.030), and cerebral infarction (correlation=.026). 3.6% of general health indicators are associated with accelerated cognitive aging, with the three largest associations being with personal history of disease (correlation=.031), problems related to lifestyles (correlation=.025), and receiving medical care (correlation=.024). 2.9% of metabolic diseases are associated with accelerated cognitive aging, with the two associations being with non-insulin-dependent type two diabetes (correlation=.034) and disorders of lipoprotein metabolism and other lipidemias (correlation=.029). We found no disease to be associated with decelerated cognitive aging.

#### Environmental variables associated with accelerated brain aging

##### Anatomical aging

The three environmental variable categories most associated with accelerated brain anatomical aging are smoking, sleep and sun exposure. Specifically, 41.7% of smoking variables are associated with accelerated brain anatomical aging, with the three largest associations being with pack years adult smoking as proportion of life span exposed to smoking (correlation=.111), pack years of smoking (correlation=.110), and past tobacco smoking: smoked on most or all days (correlation=.086). 28.6% of sleep variables are associated with accelerated brain anatomical aging, with the two associations being with napping during the day (correlation=.036) and daytime dozing/sleeping/narcolepsy (correlation=.023). 20.0% of sun exposure variables are associated with accelerated brain anatomical aging, with the three largest associations being with facial aging: do not know (correlation=.035), facial aging: older than you are (correlation=.027), and time spent outdoors in winter (correlation=.025).

Conversely, the three environmental variable categories most associated with decelerated brain anatomical aging are smoking, physical activity and sleep. Specifically, 29.2% of smoking variables are associated with decelerated brain anatomical aging, with the three largest associations being with age started smoking in former smokers (correlation=.082), smoking status: never (correlation=.078), and past tobacco smoking: I have never smoked (correlation=.044). 28.6% of physical activity variables are associated with decelerated brain anatomical aging, with the three largest associations being with usual walking pace (correlation=.067), duration walking for pleasure (correlation=.040), and duration of heavy do-it-yourself (correlation=.039). 14.3% of sleep variables are associated with decelerated brain anatomical aging (one association: snoring; correlation=.026).

##### Cognitive aging

The three environmental variable categories most associated with accelerated brain cognitive aging are smoking, sleep and sun exposure. Specifically, 29.2% of smoking variables are associated with accelerated brain cognitive aging, with the three largest associations being with smoking status: current (correlation=.040), current tobacco smoking: yes, on most or all days (correlation=.038), and number of cigarettes currently smoked daily (correlation=.036). 28.6% of sleep variables are associated with accelerated brain cognitive aging, with the two associations being with napping during the day (correlation=.061) and daytime dozing/sleeping-narcolepsy (correlation=.046). 15.0% of sun exposure variables are associated with accelerated brain cognitive aging, with the three largest associations being with time spent outdoors in winter (correlation=.063), facial aging: about your age (correlation=.040), and time spent outdoors in summer (correlation=.034).

Conversely, the three environmental variable categories most associated with decelerated cognitive aging are electronic devices, physical activity and sleep. Specifically, 40.0% of electronic device variables are associated with decelerated cognitive aging, with the three largest associations being with playing computer games (correlation=.125), hands-free device/speakerphone use with mobile phone in the last three months (correlation=.039), and usual side of head for mobile phone use: equally left or right (correlation=.025). 34.3% of physical activity variables are associated with decelerated cognitive aging, with the three largest associations being with driving faster than motorway speed limit (correlation=.083), usual walking pace (correlation=.070), and time spent using the computer (correlation=.054). 14.3% of sleep variables are associated with decelerated cognitive aging (one association: getting up in the morning; correlation=.047).

#### Socioeconomic variables associated with accelerated brain aging

##### Anatomical aging

The two socioeconomic variable categories associated with accelerated brain anatomical aging are social support (leisure/social activities: pub or social club; correlation=.046) and household (living in a flat, maisonette or apartment: correlation=.042; renting from local authority, local council or housing association: correlation=.042).

Conversely, the three socioeconomic variable categories most associated with decelerated brain anatomical aging are education, sociodemographics and employment. Specifically, 25.0% of education variables are associated with decelerated brain anatomical aging, with the two associations being with A/AS levels or equivalent (correlation=.031) and college or university degree (correlation=.031). 14.3% of sociodemographic variables are associated with decelerated brain anatomical aging (one association: not receiving any attendance/disability/mobility allowance; correlation=.046). 13.0% of employment variables are associated with decelerated brain anatomical aging, with the three largest associations being with transport type for commuting to job workplace: car/motorcycle (correlation=.033), current employment status: in paid employment or self-employed (correlation=.030), and time employed in main current job (correlation=.026).

##### Cognitive aging

The three socioeconomic variable categories most associated with accelerated brain cognitive aging are sociodemographics, employment and education. Specifically, 28.6% of sociodemographic variables are associated with accelerated brain cognitive aging, with the two associations being with private healthcare (correlation=.027) and receiving an attendance allowance (correlation=.025). 13.0% of employment variables are associated with accelerated brain cognitive aging, with the three largest associations being with jobs involving heavy manual or physical work (correlation=.039), night shift work (correlation=.036), and job involving mainly walking and standing (correlation=.030). 12.5% of education variables are associated with accelerated brain cognitive aging (one association: no qualification among the ones listed; correlation=.071).

Conversely, the three socioeconomic variable categories most associated with decelerated brain cognitive aging are education, sociodemographics and social support. Specifically, 37.5% of sociodemographic variables are associated with decelerated brain cognitive aging, with the three largest associations being with having a college or university degree (correlation=.078), A/AS levels or equivalent (correlation=.061), and O levels/GCSEs or equivalent (correlation=.051). 14.3% of sociodemographic variables are associated with decelerated brain cognitive aging (one association: not receiving attendance, disability or mobility allowance; correlation=.039). 11.1% of social support variables are associated with decelerated brain cognitive aging (one association: attending a sports club or a gym for leisure/social activities; correlation=.031).

### Correlation between accelerated brain anatomical and cognitive aging in terms of associations with biomarkers, clinical phenotypes, diseases, environmental and socioeconomic variables

We compared accelerated brain anatomical aging and cognitive aging in terms of XWAS associations. For example, in terms of environmental variables, are the diets that protect against cognitive aging the same as the diets that protect against brain anatomical aging?

We found that the correlation between accelerated brain anatomical and cognitive aging is .483 in terms of biomarkers, .657 in terms of associated clinical phenotypes, .300 in terms of diseases, .455 in terms of environmental variables and .252 in terms of socioeconomic variables (Figure 6). These results can be interactively explored at https://www.multidimensionality-of-aging.net/correlation_between_aging_dimensions/xwas_univariate.

**Figure 6:**
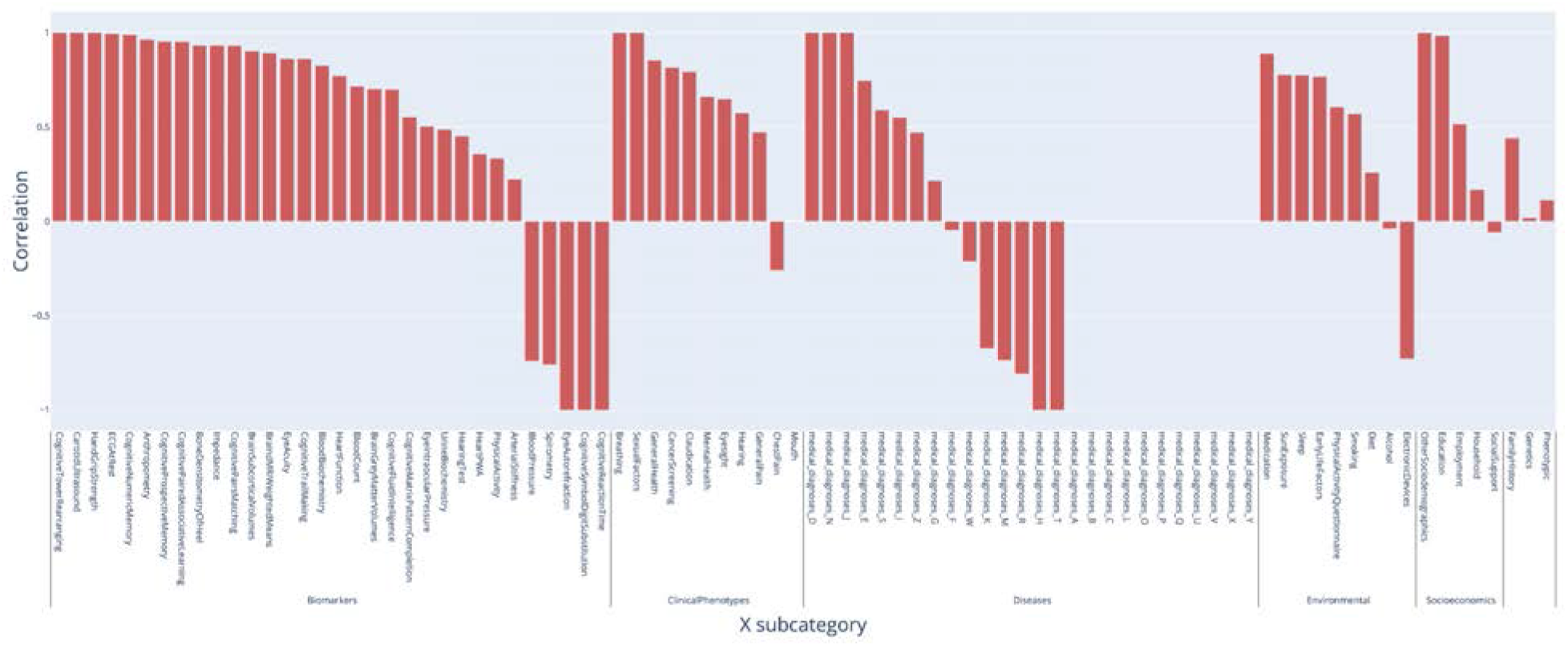
**Correlation between accelerated brain anatomical and cognitive aging in terms of associated biomarkers, clinical phenotypes, diseases, family history, environmental and socioeconomic variables**

### Predicting accelerated brain aging from biomarkers, clinical phenotypes, diseases, environmental variables and socioeconomic variables

We predicted accelerated brain aging using variables from the different X-datasets categories (biomarkers, clinical phenotypes, diseases, environmental variables and socioeconomic variables). Specifically we built a model using the variables from each of their respective subcategories (e.g blood pressure biomarkers), and found that no dataset could explain more than 5% of the variance in accelerated brain aging. This held true for brain biomarkers, as well. Cognitive biomarkers did not predict accelerated brain anatomical aging, and reciprocally.

## Discussion

### Brain anatomical and cognitive aging are two distinct phenotypes

We predicted age from brain MRI (R^2^=75.3±0.8%; RMSE=3.73±0.06 years) and cognitive tests (37.9±1.1%; RMSE=5.95±0.05 years). Brain anatomical and cognitive aging are two distinct phenotypes, with accelerated aging in these two brain dimensions being only .112±.006 phenotypically correlated and genetically uncorrelated (correlation=.019±.056).

The weak correlation between brain anatomical and cognitive aging is reflected in the XWAS results. Accelerated brain anatomical aging is associated with cognitive biomarkers, and the other way around. However, cognitive test biomarkers (e.g biomarkers for fluid intelligence) could only explain 1-2% of the variance in accelerated brain anatomical aging. Reciprocally, brain MRI biomarkers (e.g subcortical volumes) could only explain approximately 5% of the variance in accelerated cognitive aging.

### Genetic and non-genetic factors associated with accelerated brain aging

The GWASs highlighted genes associated with brain anatomy and function, confirming the clinical significance of our brain age predictors and are apparently distinct from recent GWASs on brain MRI derived phenotypes ^28^. Specifically, the GWAS performed on accelerated anatomical brain aging highlighted CRHR1 (hypothalamic-pituitary-adrenal pathway), RP11-669E14.4 (linked to Alzheimer’s disease), LRRC37A2 (linked to progressive myoclonic epilepsy-6 and Koolen-De Vries syndrome, a developmental disorder), KANSL1 (linked to Koolen-De Vries Syndrome and already associated with Alzheimer’s disease ^29^), MAPT (linked to diverse neurodegenerative disorders such as frontotemporal dementia, progressive supranuclear palsy, Alzheimer’s disease, Pick’s disease and cortico-basal degeneration) and MAPT-AS1 (linked to Parkinson Disease and Koolen-De Vries Syndrome). For accelerated cognitive aging the GWAS highlighted C6orf136 (linked to Central Nervous System Germ Cell Tumor and Spinocerebellar Ataxia). Elucidating the function of these genes and genetic loci may shed light on neurodegeneration (and/or regression) in age-related cognitive diseases.

Accelerated brain aging as defined by our predictors is associated with biomarkers, clinical phenotypes and diseases linked to brain health, such as cognitive and brain MRI biomarkers, mental health clinical phenotypes, mental disorders and intracranial injury, further confirming its biological significance. We found that accelerated brain aging is also associated with cardiovascular health (blood pressure, arterial stiffness, electrocardiogram, heart function, chest pain, heart disease), lung function (spirometry, breathing, chronic obstructive pulmonary disease), metabolic health (metabolomic biochemistry, diabetes), as well as other organ systems (e.g eyesight, hearing, blood count, physical activity), suggesting that brain aging is connected to the aging of other organ systems. Additionally, brain aging and facial aging are correlated (correlation=.058). The connection between brain aging and cardiovascular or metabolic health is already well known ^30^. More generally, brain age seems to be connected to general health (e.g overall health rating, history of disease or medical treatment). We explore the connection between brain aging and other organ systems aging more in depth in a different paper ^31^.

Reflecting the literature, accelerated aging is linked to environmental factors such as smoking ^32^, sleep ^33^, alcohol intake (we report mixed associations, partially aligned with the literature ^34^), diet^35^ and physical activity ^36^. Decelerated brain aging is correlated with socioeconomic status (education, social support, household, employment, other sociodemographics), which is likely mediated by improved access to healthcare and health literacy ^37^. The correlation between socioeconomic status and expected life expectancy is already clearly established, with the richest 1% US males living 14.6±0.2 years longer than their poorest 1% counterparts on average. Similarly, the richest 1% US females live on average 10.1±0.2 years longer than the poorest 1% females. ^38^

### Brain anatomical and cognitive features driving age prediction

We found that attention maps highlighted diverse regions of the brain, which is consistent with the systemic brain changes that happen with aging (atrophy, white matter hyperintensities, impaired white matter integrity, microbleeds, small vessel changes) ^1^ and the fact that the scalar predictors’ feature importance was distributed between the different features, with most features being considered important by at least one of the algorithms. This overall suggests that no specific part of the brain is more significantly affected by aging than the others. The ensemble model encompassing all MRI data modalities (images and scalars) significantly outperformed the ensemble model that relied exclusively on scalar features (R^2^=75.3±0.8% vs. 64.7±1.0%), suggesting that the convolutional neural networks were able to extract features from the raw images that were not extracted as scalar features by UKB.

Our finding that brain cognitive aging is associated with reduced processing speed and fluid intelligence is coherent with the literature ^2, 3^. Age-related changes in processing speed are linked to changes in the cerebellar and the prefrontal cortex ^26^ and we therefore expected that accelerated brain cognitive aging and accelerated brain anatomical aging would be more strongly correlated (correlation: .113±.007).

For both cognitive and MRI scalar-based models, non-linear algorithms (GBM, neural network) only marginally outperformed linear models (cognitive tests: R^2^=37.8±1.2% vs. 34.0±1.4%; MRI features: R^2^=62.7±1.7% vs. 62.2±1.1%), suggesting that the more complex algorithms did not find non-linearities or complex interactions between the features to exploit and improve the prediction.

### Comparison between our brain predictors and the literature

A summary of the comparison between our models and the models reported in the literature can be found in Table S24.

#### Cognitive tests

Anatürk et al. used XGBoost ^25^ on cognitive tests collected on 537 participants with a mean age of 69±5 years to predict chronological age with a R^2^ value of 9% and a RMSE of 4.77 years ^39^. Using UKB’s cognitive test data we predicted chronological age with a R^2^ value of 37.9±0.4% and a RMSE of 5.95±0.02 years. Our R^2^ value is four times higher than the one reported by Anatürk et al., which is likely driven by UKB’s sample size, which is approximately six times larger than the one reported by Anatürk et al. (UKB: 31,720; Anatürk et al.: 537). The lower RMSE reported by Anatürk et al. can be explained by the narrower age range covered by their dataset, with a standard deviation of five years.

#### Brain MRIs

A large number of models have been built to predict chronological age using brain MRI images ^6–21^. For comparison, we chose two that used three-dimensional CNN architectures to compare prediction performance. The first was built by Cole et al. on 2,001 samples aged between 18 and 90 years old and yielded a R^2^ value of 92% and a RMSE of 5.31 years ^7^. In contrast, our ensemble model for a population between ages 37-82 years gave a R^2^ value of 76.4±0.3% and a RMSE of 3.73±0.01 years. Our lower R^2^ value suggests that Cole et al.’s model outperformed our model, while our lower RMSE value suggests the opposite. This apparent contradiction can be explained by the significantly larger age range covered by Cole et al.’s dataset (18-90 years) (see Discussion - Performances - Comparison between our models and the literature in terms of prediction accuracy - Recommended considerations when comparing the prediction performances between different datasets - R^2^ vs. RMSE).

A fairer comparison can be made between our model and the model built by Jonsson et al. on the same cohort (12,395 UKB samples aged 46-79 years). Their three-dimensional (3D) CNN architecture gave a MAE of 3.63 years and a R^2^ value of 61.4% after being re-trained on the UKB dataset ^8^. This R^2^ value is significantly lower than the one we obtained with our ensemble model (R^2^=76.4±0.3%). This is perhaps surprising because, unlike Jonsson et al.’s model, our model is based on three two-dimensional sliced images of the brain in the three anatomical planes. Even without ensembling, we found that our 2D models built on the three brain anatomical planes outperformed the model reported by Jonsson et al. (Sagittal plane: R^2^=61.9±0.3%, Coronal plane: R^2^=62.6±0.3%, Transverse plane: R^2^=64.3±0.3%). One plausible explanation is that relying on 2D images allowed us to leverage deeper and more complex architectures. Additionally, Jonsson et al. reported two SNPs associated with accelerated brain aging (rs1452628 and rs2435204). We did not find an association between our accelerated brain aging phenotypes and these SNPs, which could be explained by the fact that the brain region driving this association might not be captured by our 2D brain slice images, by the difference in sample sizes, or by the difference in selection of potential confounding factors in their GWAS (Jonsson et al. corrected for total intracranial volume and head motion in addition to age and sex).

#### Brain electroencephalography

Brain electroencephalography [EEG] signals were used to predict chronological age ^23, 24^. Sun et al. engineered features from EEGs collected from 4,506 participants aged 18-80 years and used a linear regression to predict chronological age with a R^2^ value of 67.24% and a MAE of 7.6 years ^24^. Zoubi et al. engineered features from EEGs collected from 468 participants aged 18-58 years and used a support vector regression [SVR] with radial kernel to predict chronological age with a R^2^ value of 37±6% and a RMSE of 8.46±0.59 ^23^.

### Limitations

To limit the computational requirements of our analysis, we extracted two-dimensional images from the three-dimensional videos. Leveraging the raw data with a four-dimensional CNN or a three-dimensional CNN-LSTM could improve the prediction accuracy.

UKB is an observational dataset. As a consequence, the correlations that we report in the XWAS do not imply causation. In particular, many associations might be confounded by socioeconomic status. To overcome this limitation, a future direction for this project could be to leverage the subset of participants with repeated measurements. We could then for example test if specific changes in diet are associated with a reduction in biological age and correct for socioeconomic status, taking us one step closer to causality.

### Utility of brain age predictors

In conclusion, our predictors can be used to assess different facets of an individual’s brain aging process. Our results suggest potential lifestyle and therapeutic interventions to slow or reverse brain aging. Finally, our brain age predictors could be used to assess the efficiency of emerging rejuvenating ^40^ on the brain function and anatomy. Biological age predictors such as the DNA methylation clock ^41, 42^ are already being used in clinical trials ^43^, but the multidimensionality of aging ^31, 44^ may allow a more specific model to survey the effect of therapeutics on the brain.

## Methods

### Data and materials availability

We used the UK Biobank (project ID: 52887). The code can be found at https://github.com/Deep-Learning-and-Aging. The results can be interactively and extensively explored at https://www.multidimensionality-of-aging.net/. We will make the biological age phenotypes available through UK Biobank upon publication. The GWAS results can be found at https://www.dropbox.com/s/59e9ojl3wu8qie9/Multidimensionality_of_aging-GWAS_results.zip?dl=0.

### Software

Our code can be found at https://github.com/Deep-Learning-and-Aging. For the genetics analysis, we used the BOLT-LMM ^45, 46^ and BOLT-REML ^47^ softwares. We coded the parallel submission of the jobs in Bash ^48^.

### Cohort Dataset: Participants of the UK Biobank

We leveraged the UK Biobank^27^ cohort (project ID: 52887). The UKB cohort consists of data originating from a large biobank collected from 502,211 de-identified participants in the United Kingdom that were aged between 37 years and 74 years at enrollment (starting in 2006). Out of these participants, 44,183 had brain MRI collected from them. Different subsets of the 502,211 participants also took diverse cognitive tests (Figure 1A). The Harvard internal review board (IRB) deemed the research as non-human subjects research (IRB: IRB16-2145).

### Data types and Preprocessing

The data preprocessing step is different for the different data modalities: demographic variables, scalar predictors and images. We define scalar predictors as predictors whose information can be encoded in a single number, such as the fluid intelligence score, as opposed to data with a higher number of dimensions such as images (two dimensions, which are the height and the width of the image).

#### Demographic variables

First, we removed out the UKB samples for which age or sex was missing. For sex, we used the genetic sex when available, and the self-reported sex when genetic sex was not available. We computed age as the difference between the date when the patient attended the assessment center and the year and month of birth of the patient to estimate the patient’s age with greater precision. We one-hot encoded ethnicity.

#### Scalar biomarkers: anthropometry, impedance, heel bone densitometry and hand grip strength

We define scalar data as a variable that is encoded as a single number, such as the time to complete a test or the number of errors committed while solving it, as opposed to data with a higher number of dimensions, such as images. The complete list of scalar biomarkers can be found in Table S6 under “Brain”. We did not preprocess the scalar data, aside from the normalization that is described under cross-validation further below.

#### Brain Magnetic Resonance Images

The UKB contains 42,098 resting functional 3D brain MRIs videos, covering 40,597 participants (field 20227). The dimension of the files are 88*88*64 pixels, with 490 time points collected over six minutes. A “reference scan” is also available. The reference scan is a single 3D image (88*88*64 pixels) that has higher between-tissue contrast to noise and is used to correct head motion and alignment during the procedure. We selected the reference scan as well as the median time frame of the raw data (time step 245) and generated one image for each anatomical plane (sagittal, coronal, transverse). For each plane, we selected the median slice, as well as the slices that were 5 pixels before and after the median slice and saved the resulting three slices as a three-dimensional RGB image .jpeg image. We padded the images with zeros to center the brain pictures, with a resulting shape of 88*88*3 for all images. Finally, we resized the images to be 316*316*3. A sample of preprocessed brain MRI images can be found in Fig. S2.

#### Data augmentation

To prevent overfitting and increase our sample size during the training we used data augmentation ^49^ on the images. Each image was randomly shifted vertically ([-10%; +10%]) and horizontally ([-5%; +5%]), as well as rotated ([-10 degrees; 10 +degrees]). We chose the hyperparameters for these transformations’ distributions to represent the variations we observed between the images in the initial dataset. The data augmentation process is dynamically performed during the training. Augmented images are not generated in advance. Instead, each image is randomly augmented before being fed to the neural network for each epoch during the training.

### Machine learning algorithms

For scalar datasets, we used elastic nets, gradient boosted machines [GBMs] and fully connected neural networks. For images we used two-dimensional convolutional neural networks.

#### Scalar data

We used three different algorithms to predict age from scalar data (non-dimensional variables, such as laboratory values). Elastic Nets [EN] (a regularized linear regression that represents a compromise between ridge regularization and LASSO regularization), Gradient Boosted Machines [GBM] (LightGBM implementation ^50^), and Neural Networks [NN]. The choice of these three algorithms represents a compromise between interpretability and performance. Linear regressions and their regularized forms (LASSO ^51^, ridge ^52^, elastic net ^53^) are highly interpretable using the regression coefficients but are poorly suited to leverage non-linear relationships or interactions between the features and therefore tend to underperform compared to the other algorithms. In contrast, neural networks ^54, 55^ are complex models, which are designed to capture non-linear relationships and interactions between the variables. However, tools to interpret them are limited ^56^ so they are closer to a “black box”. Tree-based methods such as random forests ^57^, gradient boosted machines ^58^ or XGBoost ^25^ represent a compromise between linear regressions and neural networks in terms of interpretability. They tend to perform similarly to neural networks when limited data is available, and the feature importances can still be used to identify which predictors played an important role in generating the predictions. However, unlike linear regression, feature importances are always non-negative values, so one cannot interpret whether a predictor is associated with older or younger age. We also performed preliminary analyses with other tree-based algorithms, such as random forests ^57^, vanilla gradient boosted machines ^58^ and XGBoost ^25^. We found that they performed similarly to LightGBM, so we only used this last algorithm as a representative for tree-based algorithms in our final calculations.

### Brain MRI

#### Convolutional Neural Networks Architectures

We used transfer learning ^59–61^ to leverage two different convolutional neural networks ^62^ [CNN] architectures pre-trained on the ImageNet dataset ^63–65^ and made available through the python Keras library ^66^: InceptionV3 ^67^ and InceptionResNetV2 ^68^. We considered other architectures such as VGG16 ^69^, VGG19 ^69^ and EfficientNetB7 ^70^, but found that they performed poorly and inconsistently on our datasets during our preliminary analysis and we therefore did not train them in the final pipeline. For each architecture, we removed the top layers initially used to predict the 1,000 different ImageNet images categories. We refer to this truncated model as the “base CNN architecture”.

We added to the base CNN architecture what we refer to as a “side neural network”. A side neural network is a single fully connected layer of 16 nodes, taking the sex and the ethnicity variables of the participant as input. The output of this small side neural network was concatenated to the output of the base CNN architecture described above. This architecture allowed the model to consider the features extracted by the base CNN architecture in the context of the sex and ethnicity variables. For example, the presence of the same anatomical feature could be interpreted by the algorithm differently for a male and for a female. We added several sequential fully connected dense layers after the concatenation of the outputs of the CNN architecture and the side neural architecture. The number and size of these layers were set as hyperparameters. We used ReLU ^71^ as the activation function for the dense layers we added, and we regularized them with a combination of weight decay ^72, 73^ and dropout ^74^, both of which were also set as hyperparameters. Finally, we added a dense layer with a single node and linear activation to predict age.

#### Compiler

The compiler uses gradient descent ^75, 76^ to train the model. We treated the gradient descent optimizer, the initial learning rate and the batch size as hyperparameters. We used mean squared error [MSE] as the loss function, root mean squared error [RMSE] as the metric and we clipped the norm of the gradient so that it could not be higher than 1.0 ^77^.

We defined an epoch to be 32,768 images. If the training loss did not decrease for seven consecutive epochs, the learning rate was divided by two. This is theoretically redundant with the features of optimizers such as Adam, but we found that enforcing this manual decrease of the learning rate was sometimes beneficial. During training, after each image has been seen once by the model, the order of the images is shuffled. At the end of each epoch, if the validation performance improved, the model’s weights were saved.

We defined convergence as the absence of improvement on the validation loss for 15 consecutive epochs. This strategy is called early stopping ^78^ and is a form of regularization. We requested the GPUs on the supercomputer for ten hours. If a model did not converge within this time and improved its performance at least once during the ten hours period, another GPU was later requested to reiterate the training, starting from the model’s last best weights.

### Training, tuning and predictions

We split the entire dataset into ten data folds. We then tuned the models built on scalar data and the models built on images using two different pipelines. For scalar data-based models, we performed a nested-cross validation. For images-based models, we manually tuned some of the hyperparameters before performing a simple cross-validation. We describe the splitting of the data into different folds and the tuning procedures in greater detail in the Supplementary.

### Interpretability of the machine learning predictions

To interpret the models, we used the regression coefficients for the elastic nets, the feature importances for the GBMs, a permutation test for the fully connected neural networks, and attention maps (saliency and Grad-RAM) for the convolutional neural networks (Supplementary Methods).

### Ensembling to improve prediction and define aging dimensions

We built a three-levels hierarchy of ensemble models to improve prediction accuracies. At the lowest level, we combined the predictions from different algorithms on the same aging sub-subdimension. For example, we combined the predictions generated by the elastic net, the gradient boosted machine and the neural network on the cognitive test (dimension) “TowerRearranging” (subdimension). At the second lowest level, we combined the predictions from different subdimensions of a unique dimension. For example, we combined the predictions from the different cognitive tests (e.g. ReactionTime, MatrixPatternCompletion and TowerRearranging) into an ensemble prediction for the “Cognitive” subdimension. Finally, at the highest level, we combined the predictions from anatomical and cognitive brain features into a general brain age prediction. The ensemble models from the lower levels are hierarchically used as components of the ensemble models of the higher models. For example, the ensemble model built by combining the algorithms trained on fluid intelligence variables is leveraged when building the general brain aging ensemble model.

We built each ensemble model separately on each of the ten data folds. For example, to build the ensemble model on the testing predictions of the data fold #1, we trained and tuned an elastic net on the validation predictions from the data fold #0 using a 10-folds inner cross-validation, as the validation predictions on fold #0 and the testing predictions on fold #1 are generated by the same model (see Methods - Training, tuning and predictions - Images - Scalar data - Nested cross-validation; Methods - Training, tuning and predictions - Images - Cross-validation). We used the same hyperparameters space and Bayesian hyperparameters optimization method as we did for the inner cross-validation we performed during the tuning of the non-ensemble models.

To summarize, the testing ensemble predictions are computed by concatenating the testing predictions generated by ten different elastic nets, each of which was trained and tuned using a 10-folds inner cross-validation on one validation data fold (10% of the full dataset) and tested on one testing fold. This is different from the inner-cross validation performed when training the non-ensemble models, which was performed on the “training+validation” data folds, so on 9 data folds (90% of the dataset).

### Evaluating the performance of models

We evaluated the performance of the models using two different metrics: R-Squared [R^2^] and root mean squared error [RMSE]. We computed a confidence interval on the performance metrics in two different ways. First, we computed the standard deviation between the different data folds. The test predictions on each of the ten data folds are generated by ten different models, so this measure of standard deviation captures both model variability and the variability in prediction accuracy between samples. Second, we computed the standard deviation by bootstrapping the computation of the performance metrics 1,000 times. This second measure of variation does not capture model variability but evaluates the variance in the prediction accuracy between samples.

### Brain age definition

We defined the biological age of participants as the prediction generated by the model corresponding to brain dimension or subdimension, after correcting for the bias in the residuals.

We indeed observed a bias in the residuals. For each model, participants on the older end of the chronological age distribution tend to be predicted younger than they are. Symmetrically, participants on the younger end of the chronological age distribution tend to be predicted older than they are. This bias does not seem to be biologically driven. Rather it seems to be statistically driven, as the same 60-year-old individual will tend to be predicted younger in a cohort with an age range of 60-80 years, and to be predicted older in a cohort with an age range of 60-80. We ran a linear regression on the residuals as a function of age for each model and used it to correct each prediction for this statistical bias.

After defining biological age as the corrected prediction, we defined accelerated aging as the corrected residuals. For example, a 60-year-old whose cognitive tests predicted an age of 70 years old after correction for the bias in the residuals is estimated to have a cognitive age of 70 years, and an accelerated cognitive aging of ten years.

It is important to understand that this step of correction of the predictions and the residuals takes place after the evaluation of the performance of the models but precedes the analysis of the brain ages properties.

### Genome-wide association study of accelerated brain aging

The UKB contains genome-wide genetic data for 488,251 of the 502,492 participants^79^ under the hg19/GRCh37 build.

We used the average accelerated aging value over the different samples collected over time (see Supplementary - Generating average predictions for each participant). Next, we performed genome wide association studies [GWASs] to identify single-nucleotide polymorphisms [SNPs] associated with accelerated aging in each brain dimension using BOLT-LMM ^45, 46^ and estimated the the SNP-based heritability for each of our biological age phenotypes, and we computed the genetic pairwise correlations between dimensions using BOLT-REML ^47^. We used the v3 imputed genetic data to increase the power of the GWAS, and we corrected all of them for the following covariates: age, sex, ethnicity, the assessment center that the participant attended when their DNA was collected, and the 20 genetic principal components precomputed by the UKB. We used the linkage disequilibrium [LD] scores from the 1,000 Human Genomes Project^80^. To avoid population stratification, we performed our GWAS on individuals with White ethnicity.

#### Identification of SNPs associated with accelerated brain aging

We identified the SNPs associated with accelerated brain aging dimensions using the BOLT-LMM ^45, 46^ software (p-value of 5e-8). The sample size for the genotyping of the X chromosome is one thousand samples smaller than for the autosomal chromosomes. We therefore performed two GWASs for each aging dimension. (1) excluding the X chromosome, to leverage the full autosomal sample size when identifying the SNPs on the autosome. (2) including the X chromosome, to identify the SNPs on this sex chromosome. We then concatenated the results from the two GWASs to cover the entire genome, at the exception of the Y chromosome.

We plotted the results using a Manhattan plot and a volcano plot. We used the bioinfokit ^81^ python package to generate the Manhattan plots. We generated quantile-quantile plots [Q-Q plots] to estimate the p-value inflation as well.

#### Heritability and genetic correlation

We estimated the heritability of the accelerated aging dimensions using the BOLT-REML ^47^ software. We included the X chromosome in the analysis and corrected for the same covariates as we did for the GWAS. Using the same software and parameters, we computed the genetic correlations between accelerated aging in the different brain dimensions.

We annotated the significant SNPs with their matching genes using the following four steps pipeline. (1) We annotated the SNPs based on the rs number using SNPnexus ^82–86^. When the SNP was between two genes, we annotated it with the nearest gene. (2) We used SNPnexus to annotate the SNPs that did not match during the first step, this time using their genomic coordinates. After these two first steps, 30 out of the 9,697 significant SNPs did not find a match. (3) We annotated these SNPs using LocusZoom ^87^. Unlike SNPnexus, LocusZoom does not provide the gene types, so we filled this information with GeneCards ^88^. After this third step, four genes were not matched. (4) We used RCSB Protein Data Bank ^89^ to annotate three of the four missing genes.

### Non-genetic correlates of accelerated aging

We identified non-genetically measured (i.e factors not measured on a GWAS array) correlates of each brain dimension, which we classified in six categories: biomarkers, clinical phenotypes, diseases, family history, environmental, and socioeconomic variables. We refer to the union of these association analyses as an X-Wide Association Study [XWAS]. (1) We define as biomarkers the scalar variables measured on the participant, which we initially leveraged to predict age (e.g. blood pressure, Table S6). (2) We define clinical phenotypes as other biological factors not directly measured on the participant, but instead collected by the questionnaire, and which we did not use to predict chronological age. For example, one of the clinical phenotypes categories is eyesight, which contains variables such as “wears glasses or contact lenses”, which is different from the direct refractive error measurements performed on the patients, which are considered “biomarkers” (Table S9). (3) Diseases include the different medical diagnoses categories listed by UKB (Table S12). (4) Family history variables include illnesses of family members (Table S15). (5) Environmental variables include alcohol, diet, electronic devices, medication, sun exposure, early life factors, medication, sun exposure, sleep, smoking, and physical activity variables collected from the questionnaire (Table S18). (6) Socioeconomic variables include education, employment, household, social support and other sociodemographics (Table S21). We provide information about the preprocessing of the XWAS in the Supplementary Methods.

## Data Availability

https://www.multidimensionality-of-aging.net/

https://www.dropbox.com/s/59e9ojl3wu8qie9/Multidimensionality_of_aging-GWAS_results.zip?dl=0

## Author Contributions

**Alan Le Goallec:** (1) Designed the project. (2) Supervised the project. (3) Predicted chronological age from images. (4) Computed the attention maps for the images. (5) Ensembled the models, evaluated their performance, computed biological ages and estimated the correlation structure between the brain aging dimensions. (6) Performed the genome wide association studies. (5) Designed the website. (6) Wrote the manuscript.

**Samuel Diai:** (1) Predicted chronological age from the scalar features. (2) Coded the algorithm to obtain balanced data folds across the different datasets. (3) Wrote the python class to build an ensemble model using a cross-validated elastic net. (4) Performed the X-wide association study. (5) Implemented a first version of the website https://www.multidimensionality-of-aging.net/.

**Sasha Collin:** (1) Preprocessed the brain magnetic resonance images.

**Théo Vincent:** (1) Website data engineer. (2) Implemented a second version of the website https://www.multidimensionality-of-aging.net/.

**Chirag J. Patel:** (1) Supervised the project. (2) Edited the manuscript. (3) Provided funding.

## Acknowledgments

We would like to thank Raffaele Potami from Harvard Medical School research computing group for helping us utilize O2’s computing resources. We thank HMS RC for computing support. We also want to acknowledge UK Biobank for providing us with access to the data they collected. The UK Biobank project number is 52887.

## Funding

NIEHS R00 ES023504

NIEHS R21 ES25052.

NIAID R01 AI127250

NSF 163870

MassCATS, Massachusetts Life Science Center Sanofi

The funders had no role in the study design or drafting of the manuscript(s).

